# Growth, physical and cognitive function in children who are born HIV-free: school-age follow-up of a cluster-randomized trial in rural Zimbabwe

**DOI:** 10.1101/2024.01.15.24301305

**Authors:** Joe D Piper, Clever Mazhanga, Marian Mwapaura, Gloria Mapako, Idah Mapurisa, Tsitsi Mashedze, Eunice Munyama, Maria Kuona, Thombizodwa Mashiri, Kundai Sibanda, Dzidzai Matemavi, Monica Tichagwa, Soneni Nyoni, Asinje Saidi, Manasa Mangwende, Dzivaidzo Chidhanguro, Eddington Mpofu, Joice Tome, Gabriel Mbewe, Batsirai Mutasa, Bernard Chasekwa, Handrea Njovo, Chandiwana Nyachowe, Mary Muchekeza, Kuda Mutasa, Virginia Sauramba, Ceri Evans, Melissa J Gladstone, Jonathan C Wells, Elizabeth Allen, Melanie Smuk, Jean H Humphrey, Lisa F Langhaug, Naume V Tavengwa, Robert Ntozini, Andrew J Prendergast

## Abstract

**Background:** Globally, over 16 million children were exposed to HIV during pregnancy but remain HIV-free at birth and throughout childhood. Children born HIV-free (CBHF) have higher morbidity and mortality and poorer neurodevelopment in early life compared to children who are HIV-unexposed (CHU), but long-term outcomes remain uncertain. We characterized school-age growth, cognitive and physical function in CBHF and CHU previously enrolled in the Sanitation Hygiene Infant Nutrition Efficacy (SHINE) trial in rural Zimbabwe.

**Methods and Findings:** Children in SHINE who had been followed to age 18 months, were re-enrolled to this follow-up study if they were aged 7 years, resident in Shurugwi district, and had a known pregnancy HIV exposure status. From 5280 pregnant women originally enrolled, 264 CBHF and 990 CHU were evaluated at age 7 years using the School-Age Health, Activity, Resilience, Anthropometry and Neurocognitive (SAHARAN) toolbox. Cognitive function was evaluated using the Kaufman Assessment Battery for Children (KABC-II), with additional tools measuring executive function, literacy, numeracy, fine motor skills and socioemotional function. Physical function was assessed using standing broad jump and handgrip for strength, and the shuttle-run test for cardiovascular fitness. Growth was assessed by anthropometry. Body composition was assessed by bioimpedance analysis for lean mass and skinfold thicknesses for fat mass. A caregiver questionnaire measured demographics, socioeconomic status, nurturing, child discipline, food and water insecurity. We prespecified the primary comparisons and used generalized estimating equations (GEE) with an exchangeable working correlation structure to account for clustering. Adjusted models used covariates derived from directed acyclic graphs, with separate models adjusted for contemporary and early-life variables. We found strong evidence that cognitive function was lower for CBHF compared to CHU across multiple domains. The adjusted mean difference in the mental processing index (MPI), derived from KABC-II, was 3 points lower (95%CI 2, 4; P<0.001) in CBHF versus CHU. Similarly, the school achievement test (SAT) of literacy and numeracy was 7 points lower (95%CI 4, 11, P<0.001) and executive function, measured by the Plus-EF tablet-based test, was 5 points lower (95%CI 2, 8; P<0.001) in CBHF compared to CHU. CBHF also had smaller head circumferences by 0.3cm (95%CI 0.1, 0.5; P=0.009). CBHF had fewer years of schooling exposure and caregiver schooling, with higher rates of caregiver depression. CBHF had lower cardiovascular fitness from the shuttle-run test with a maximal oxygen consumption (VO_2_max) 0.8 ml/kg/min (95%CI 0.4, 1.2; P<0.001) lower than CHU. We found no evidence of differences in other growth, body composition or physical function outcomes. The main limitation of our study is that it was restricted to one of two previous study districts, with possible survivor bias and selection bias from children who had moved away.

**Conclusions:** CBHF have reductions in cognitive function, head circumference and cardiovascular fitness compared to CHU at 7 years of age. Further research is needed to define the biological and psychosocial mechanisms underlying these differences, to inform future interventions that help CBHF thrive across the life-course.

## INTRODUCTION

Increasing coverage of prevention of mother-to-child transmission (PMTCT) interventions has advanced progress towards paediatric HIV elimination in sub-Saharan Africa. The vast majority of children born to women with HIV are therefore now HIV-free, meaning they were exposed to HIV in pregnancy but remain uninfected themselves. The current global population of children born HIV-free (CBHF) is increasing, with estimates of 14.8 million in 2018[1, 2], and 15.9 million in 2021[3]. A disparity in clinical outcomes between CBHF and children who are HIV-unexposed (CHU) emerged in the pre-antiretroviral therapy (ART) era, with 3-fold higher mortality, higher frequency and severity of common infections, and more growth failure in CBHF[4, 5]. Clinical outcomes in the PMTCT era have been uncertain due to a paucity of studies, but emerging data from sub-Saharan Africa confirm that disparities persist in early life despite high coverage of maternal ART. CBHF are more likely to be born premature and small-for-gestational age[6], have a higher frequency of stunting[7], and poorer neurodevelopment[8] than CHU by 2 years of age. Multiple factors are likely to contribute to these clinical disparities[9] including both universal and HIV-specific risk factors, and the shared mechanisms through which they operate. Early-life exposures such as maternal HIV, co-infections, dysbiosis, inflammation, malnutrition and stress may have persistent effects long after the antenatal exposure has ended.

Few studies have conducted long-term follow-up of CBHF to establish whether early-life clinical disparities persist to school age. One study in Zambia found that differences in growth between CBHF and CHU had widened by 7.5 years of age compared to infancy[10], while a study of neurocognitive outcomes across five African countries[11] found no differences between CBHF and CHU groups at median age 7 years, but did not evaluate language, which may be most predictive of future function[11]. Other cohorts have reported poorer mathematic abilities[12] and reduced IQ among CBHF in south-east Asia[13], but studies have focused on a limited range of outcomes. The Sanitation Hygiene Infant Nutrition Efficacy (SHINE) trial in rural Zimbabwe evaluated the effects of improved infant and young child feeding (IYCF) and/or improved water, sanitation and hygiene (WASH) on child stunting and anaemia at 18 months of age. The trial showed that 50% of CBHF were stunted by age 18 months, and that neurodevelopmental scores were lower at 2 years of age among CBHF compared to CHU[14, 15]. Here, we report follow-up of a subgroup of CBHF and CHU at age 7 years to evaluate whether disparities in growth, physical and cognitive function persist to school age[16]. School-age function is highly predictive of later adult function, and therefore represents an important period of mid-childhood. Understanding whether CBHF thrive less well in the long-term is critical to inform the timing of interventions to improve long-term human capital in this expanding global population.

## Methods

### SHINE trial

The design and results of the SHINE trial have been previously reported[17]. In brief, between 2012 and 2015, SHINE recruited 5280 women during pregnancy from two rural Zimbabwean districts with 15% antenatal HIV prevalence (ClinicalTrials.gov NCT01824940). Women who resided in the study area were cluster-randomized to one of four intervention arms: standard-of-care (SOC, including promotion of PMTCT and optimal breastfeeding); IYCF (20 g daily small-quantity lipid-based nutrient supplement (SQ-LNS) for their infant from 6-18 months of age, with complementary feeding counselling); WASH (ventilated improved pit latrine and two handwashing stations, monthly liquid soap and chlorine, a play-space to separate children from livestock and to reduce geophagia, plus hygiene counselling); or IYCF plus WASH (all interventions). IYCF improved linear growth and haemoglobin at 18 months in both CBHF and CHU[14, 18], while WASH had no effect. The combined IYCF+WASH intervention improved child neurodevelopment at 2 years of age among CBHF, but neither intervention improved neurodevelopment in CHU[19, 20].

### School-age follow-up

To evaluate the long-term effects of HIV exposure on child health outcomes, we designed a sub-study to assess child growth, body composition, physical and cognitive function at 7 years of age. No further trial interventions had been provided after age 18 months. The protocol and statistical analysis plan for the follow-up study are registered at https://osf.io/8e2zh. Children were eligible if they were aged 7-8 years and still resident in Shurugwi district. Children were ineligible if they were no longer resident in Shurugwi, had an unknown maternal pregnancy HIV status, or were outside the age window. Among all children born to HIV-negative mothers (CHU) and evaluated at the trial endline at age 18 months, 250 per intervention arm meeting the eligibility criteria were randomly selected by computer; those who were unable to be visited or whose family declined participation were replaced by another eligible child randomly selected from the same trial arm. For CBHF, all children in Shurugwi district who were born to HIV-positive mothers and who tested HIV-negative at 18 months were offered enrolment. Families were approached through community health workers (CHWs) to determine if the child was available, and the household was interested in the follow-up study. Written informed consent from the primary caregiver and written assent from the child were obtained by research nurses, following a demonstration of the tools used to conduct the measurements.

#### HIV status

Mothers in the SHINE cohort were tested during pregnancy using an antibody rapid test algorithm (Alere Determine HIV-1/2 test, followed by INSTI HIV-1/2 test if positive). Mothers were offered HIV testing at baseline and again at 32 weeks for those who tested negative at baseline; further testing was offered at 18 months postpartum. CHU were defined as children born to mothers testing negative for HIV during pregnancy. Children who were HIV-exposed were defined as those born to mothers testing positive for HIV during pregnancy[20]. CBHF were defined as children who were HIV-exposed and confirmed HIV-negative through 18 months of age (trial endpoint)[14]. The early-life child HIV status was determined by dried blood-spot DNA polymerase chain reaction (PCR), plasma RNA PCR, or rapid test algorithm, depending on child age and sample type.

For the school-age visit, HIV testing was offered to all caregiver-child pairs to ensure that an updated HIV exposure status (in case of maternal incident infection since the end of the trial) and infection status (in case of prolonged breastfeeding and postnatal transmission since the 18-month trial endpoint) was available. If the mother’s HIV test was negative, the child was not tested. If the mother tested positive for HIV, declined testing or was unavailable, HIV testing was offered to the child with age-appropriate assent using role plays. The Determine HIV-1/2 rapid test (Abbott) was used for initial testing; positive results were repeated using the HIV 1/2 Stat-Pak rapid test (Chembio). Any children who tested positive for HIV were referred to local clinics for ART and were excluded from this analysis. All assessments were done by study nurses who were blinded to the caregiver and child HIV status.

### School-age measurements using the SAHARAN toolbox

We developed a battery of tests to measure growth, physical and cognitive outcomes, termed the School-Age Health, Activity, Resilience, Anthropometry and Neurocognitive (SAHARAN) toolbox[21]. Measurements were performed during a single home visit by extensively trained and supervised primary care nurses, using one or two tents pitched in the household or nearby. Briefly, the SAHARAN toolbox comprises a caregiver questionnaire, child questionnaire, and direct tests undertaken with the child to assess cognitive function, growth and physical function. All tests had standardised explanations, demonstrations, and translations in local languages (Shona and Ndebele). Data collectors underwent standardisations in anthropometry and cognition tests every 9 months and had regular supported supervision.

Cognitive function was assessed using the Kaufman Assessment Battery for Children 2^nd^ edition (KABC-II; Pearson UK)[22, 23]; a previously piloted School Achievement Test (SAT) which measures literacy and numeracy; and a coordination test[24] which measures fine motor skills by finger tapping. Three subtests from the android tablet-based Plus-EF test[25] measured executive function. The child’s socioemotional function and behaviour were measured by the caregiver-reported Strengths and Difficulties Questionnaire (SDQ) (https://www.sdqinfo.com/). An overall score of the child’s functional abilities was provided by the Washington Group/UNICEF screening tool[26], which helped identify underlying physical disabilities in sight, hearing and mobility, or behavioural and learning difficulties.

Growth measurements included height, knee-heel length, weight, head circumference, mid-upper arm circumference (MUAC), and waist, hip and calf circumferences. Body composition was assessed using Holtain calipers (Crosswell, UK) to measure peripheral subcutaneous fat (triceps and calf skinfold thicknesses) and central subcutaneous fat (subscapular and supra-iliac skinfold thicknesses). Bioimpedance analysis (BIA) using a BodyStat 1500 MDD machine (BodyStat, Isle of Man, UK) assessed lean mass, measured as the impedance index (height^2^/impedance) and lean mass index (1/impedance)[27]; and tissue health, measured as the phase angle.

Physical function outcomes included; leg strength, by measuring the distance jumped in the broad jump from a standing position, and handgrip strength in each hand using a dynamometer (Takei, Japan). Cardiovascular fitness was measured by the shuttle-run test, where the child repeatedly ran between two markers placed 20 metres apart, arriving at each end before a timed beep from an android app (Beep Test, Ruval Enterprises, Canada) connected to a Bluetooth speaker. The child ran until either they missed the beep three times in a row or stopped due to tiredness. VO_2_max, which represents the maximum rate at which the body uses oxygen during exercise, was calculated[28]. Resting and post-exercise blood pressure was measured using a manual aneroid sphygmomanometer (Medisave, UK). Haemoglobin was measured (Hemocue) on a finger-prick blood sample.

A caregiver questionnaire was used to record contemporary household demographics, socioeconomic status using a locally validated wealth index[29], schooling exposure, adversities, nurturing (including the Child-Parent Relationship scale[30] and the MICS Child Discipline questionnaire[31, 32]), caregiver depression[33], gender norms[34], caregiver social support[34], and food[35, 36] and water[37] insecurity.

Most data were collected electronically using Open Data Kit (ODK, opendatakit.org) on android tablets (Samsung Galaxy Tab A). The KABC-II and SAT used paper forms subsequently transcribed onto ODK. The Plus-EF tool recorded data directly into the Plus-EF application.

### Statistical analysis

A pre-specified statistical analysis plan is available at https://osf.io/8e2zh. Stata v13 and v17 were used for analyses. Baseline characteristics between CBHF and CHU groups were compared using multinomial and ordinal regression models and Somers’ D for medians, while handling within-cluster correlation with robust variance estimation. Generalised estimating equations (GEE) were used to compare each functional outcome between CBHF and CHU groups. Model 1 adjusted for trial factors: randomized intervention arm, study nurse performing the assessment, exact age of child, sex of child, and calendar season. Model 2 included these trial factors plus contemporary socioeconomic and demographic confounders identified from a directed acyclic graph (DAG) with online software (Dagitty.net; see Supporting Information): socioeconomic status, caregiver depression (Edinburgh Postnatal Depression Score, EPDS), household food insecurity assessment scale (HFIAS), household religion, caregiver social support[38], caregiver gender norms[38], caregiver age, caregiver education, adversity score and the number of children’s books at home. Model 3 adjusted for the trial factors and baseline socioeconomic and demographic confounders: birthweight, baseline EPDS, household dietary diversity score, health facility births, maternal haemoglobin, household socioeconomic score, maternal education and maternal gender norms. A pre-specified subgroup analysis was also performed if there was evidence for an interaction of CBHF with child sex (p<0.10).

### Ethics

The Medical Research Council of Zimbabwe approved the study protocol (MRCZ/A/1675). The SHINE follow-up study was registered with the Pan-African Clinical Trials Registry (PACTR202201828512110). Written informed consent from the primary caregiver and written assent from the child were obtained, following a demonstration of the tools used to conduct the measurements.

## Results

Between 22 November 2012 and 27 March 2015, 5280 pregnant women were enrolled from 211 clusters at median 12 (IQR 9, 16) gestational weeks in Chirumanzu and Shurugwi districts (Figure 1). In Shurugwi, there were 420 births to women with HIV and 2174 births to women without HIV; 376 HIV-exposed and 2016 HIV-unexposed children completed the 18-month primary endpoint visit. Between 18 months and 7 years, 2 (0.5%) CBHF and 5 (0.2%) CHU died, whilst 6 (1.6%) CBHF and 42 (2.1%) CHU were lost to follow-up. Two caregivers of CBHF and 9 caregivers of CHU declined follow-up at 7 years. There were 87 (23.2%) relocations among CBHF and 387 (19.2%) relocations among CHU; these children were therefore ineligible for inclusion. Overall, 273 HIV-exposed children were measured, of whom 5 were HIV positive and 4 had severe disability, leaving 264 CBHF in the current analysis. Of 1002 HIV-unexposed children measured, 12 were subsequently excluded: 2 were HIV-positive (due to mothers seroconverting during breastfeeding) and 10 had severe disability; 990 CHU were included in the current analysis.

**Figure 1:**
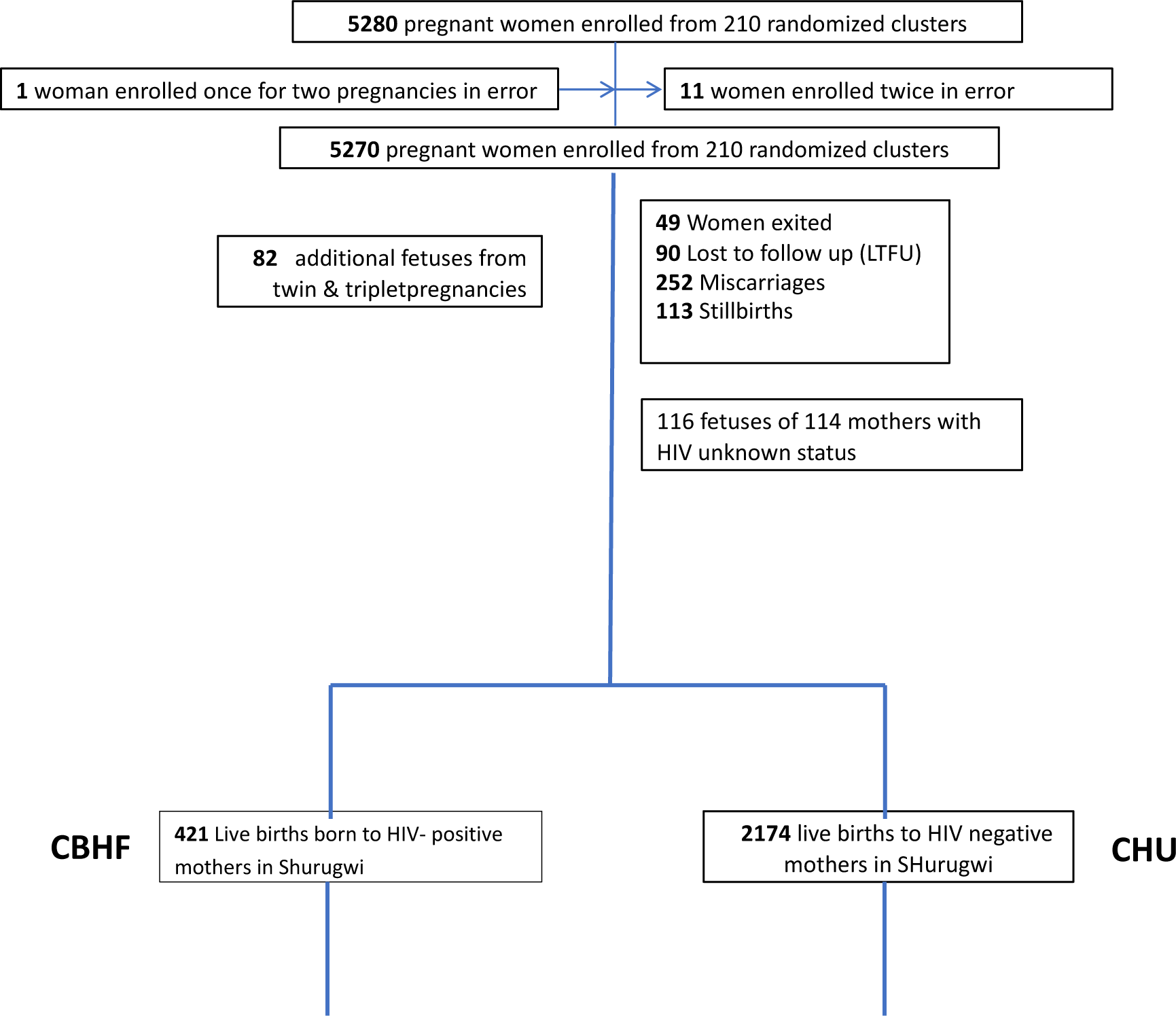
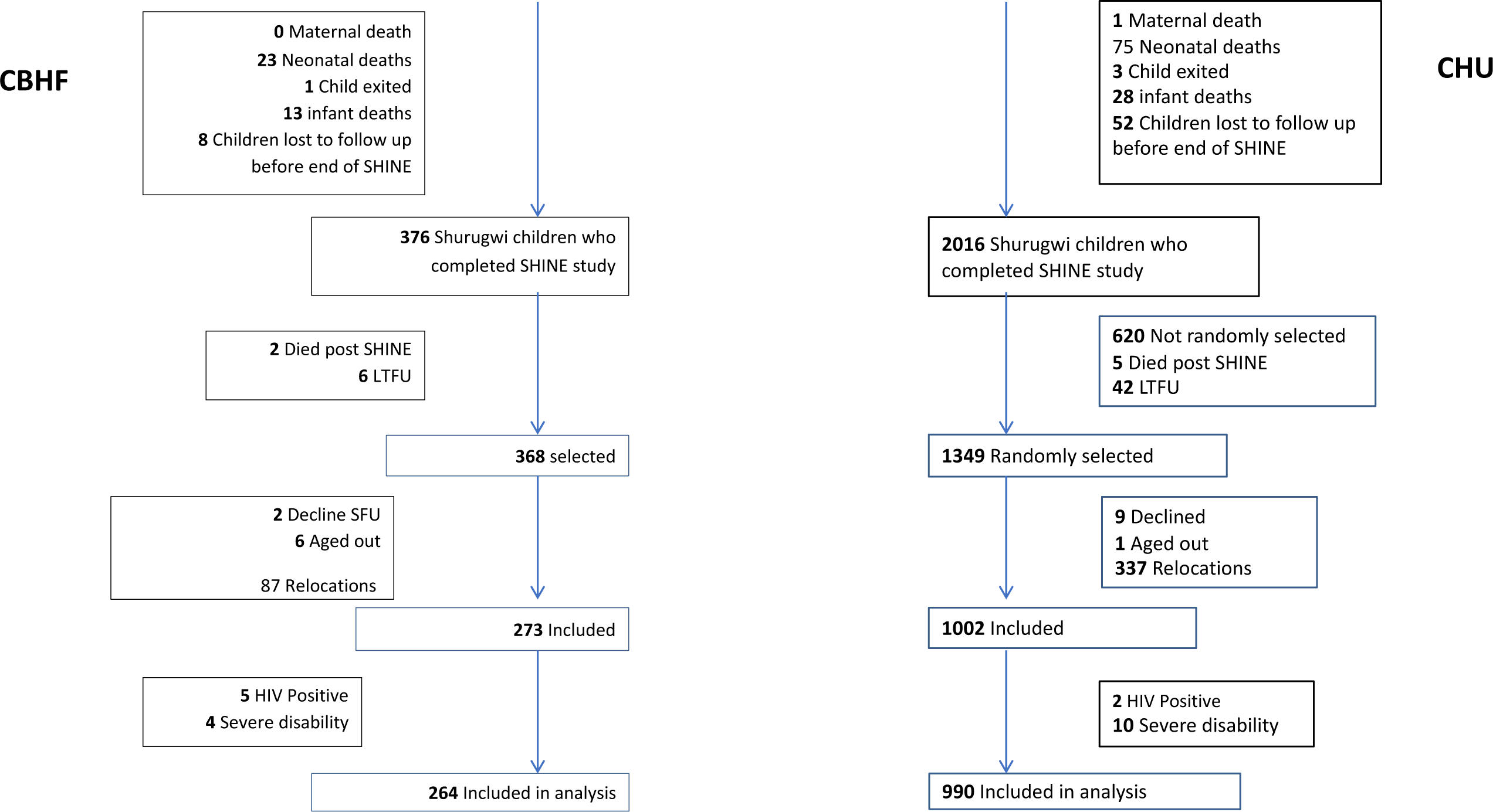
CONSORT diagram showing CBHF and CHU selected into SHINE Follow-up. Note that 6 children born to HIV positive women aged out because initially children with known HIV positive status were not included for measurement. 1 child born to HIV negative mothers also aged out due to heavy rains making their area inaccessible before they turned 8 years. Participants were recorded as lost to follow-up (LTFU) at three stages: shortly after enrolment as pregnant mothers, during the first 18 months before the trial primary endpoint was measured, or between 18 months and 7 years. Children with severe disability or who were HIV positive were not included in the current analysis.

### Participant characteristics

Characteristics of the child, caregiver and household at the time of the follow-up visit are shown in Table 1. CBHF compared to CHU had a higher proportion of caregivers who were mothers (83.1% vs 75.9%) compared to other types of caregiver such as grandmothers or other family members. CBHF also had a greater proportion had been randomized to WASH or combined arms. CBHF households had higher levels of food insecurity as measured by the HFIAS score (12.9 vs 12.0) and a higher adversity score (1.95 vs 1.77) compared to CHU households. Caregivers of CBHF versus CHU had slightly fewer years of schooling (9.5 vs 10.0), although both were relatively high. CBHF caregivers also higher depression scores (4.1 vs 3.2) compared to CHU. CBHF also had lower total schooling exposure (3.1 vs 3.3 years).

**Table 1:** Baseline characteristics. Variables all measured at the time of the follow-up visit. Baseline factors measured during participation in the original trial are shown in Supplementary Table 1. [21]. All scales are further explained in supplementary materials. IQR: inter-quartile range.

| **Variable at 7-year visit** | **CBHF**<br>**N=264** | **CHU**<br>**N=990** | **p-value** |
| --- | --- | --- | --- |
| Proportion female [N] | 51.1% [135] | 51.1% [506] | 0.99 |
| Child age, years; mean (SD) | 7.3 (0.3) | 7.3 (0.2) | 0.07 |
| Mean years of schooling (SD) | 3.1 (0.7) | 3.3 (0.8) | 0.01 |
| **Caregiver characteristics** |  |  |  |
| Mother as caregiver, % [N] | 83.3% [219] | 75.8% [750] | 0.01 |
| Mean years of schooling (SD) | 9.5 (2.7) | 10.0 (2.6) | 0.01 |
| Mean Edinburgh Postnatal Depression score (SD) | 4.0 (4.7) | 3.2 (4.4) | 0.01 |
| Mean Social support score (SD) | 3.8 (0.6) | 3.9 (0.5) | 0.38 |
| Mean Gender norm attitudes (SD) | 4.1 (0.6) | 4.1 (0.6) | 0.62 |
| Mean Total Discipline score (SD) | 2.0 (2.1) | 1.9 (2.0) | 0.62 |
| Mean Child parent relationship score (SD) | 3.3 (0.8) | 3.3 (0.7) | 0.21 |
| **Household characteristics** |  |  |  |
| SES quintile, % [N] |  |  |  |
| lowest | 23.7% [62] | 19.1% [186] | 0.50 |
| second | 20.3% [53] | 19.8% [193] |  |
| middle | 19.2% [50] | 20.2% [197] |  |
| fourth | 19.2% [51] | 20.2% [197] |  |
| highest | 17.7% [46] | 20.6% [201] |  |
| Mean SES scale (SD) | 1.5 (0.7) | 1.6 (0.6) | 0.06 |
| Mean Household Food Insecurity Assessment Scale (SD) | 12.9 (4.7) | 12.0 (4.2) | 0.005 |
| Mean Household Dietary Diversity Scale (SD) | 7.7 (2.0) | 7.7 (1.8) | 0.91 |
| Mean Total Household Water Insecurity Experiences scale (SD) | 12.2 (1.3) | 12.1 (0.9) | 0.41 |
| Female headed household, % [N] | 18.6% [50] | 17.1% [171] | 0.56 |
| Mean Adversity Score (SD) | 1.9 (1.5) | 1.8 (1.4) | 0.07 |
| Median number of children's books at home (IQR) | 0 (0, 1) | 0 (0, 1) | 0.02 |
| **Original trial arm, % [N]** |  |  |  |
| SOC | 19.3% [51] | 24.9% [246] | <0.001 |
| IYCF | 17.4% [46] | 25.3% [250] |  |
| WASH | 35.2% [93] | 25.0% [247] |  |
| Combined | 28.0% [74] | 25.0% [247] |  |

Baseline characteristics of the mother and household, which were measured at enrolment to the original trial during pregnancy, together with child characteristics between birth and 18 months (trial endpoint) are shown in Suppl Table 1. Mothers with HIV had lower haemoglobin and MUAC, and higher parity than mothers without HIV, together with higher depression and food insecurity scores, and lower socioeconomic scores. In pregnancy, women with HIV had a mean (SD) CD4 count of 474 (215); 83% were receiving ART, which was predominantly tenofovir-based regimens (70%). Between birth and 18 months, CBHF had lower anthropometry, with higher rates of stunting and a lower breastfeeding duration, compared to CHU (see Suppl Table 1).

### Cognitive, physical and growth outcomes

Cognitive outcomes for CBHF and HUU children are shown in Table 2. CBHF had lower total neurodevelopmental scores, as measured by the Mental Processing Index from the KABC-II test, which reflects overall cognitive function. There was still strong evidence of difference after adjustment for contemporary or baseline factors. CBHF also had lower scores on the School Achievement Test and showed reduced executive function as measured by the Plus-EF score. Fine motor function was lower among CBHF, with weaker evidence of difference after adjustment for contemporary or baseline factors. There was no evidence of difference between groups in the Strengths and Difficulties Questionnaire or total socioemotional score. Taken together, CBHF showed 0.2-0.3 standard deviation reductions in neurodevelopmental scores across a range of measures of cognition, executive function and academic achievement.

**Table 2:** Cognitive function in CBHF and CHU. Model 1 is adjusted for trial factors (arm, study nurse, exact child age, calendar month recruited, temperature, sex). Model 2 is adjusted for trial factors from Model 1 and contemporary factors (socioeconomic status, caregiver depression score (EPDS), household food insecurity (HFIAS), household religion, caregiver social support, caregiver gender norms, caregiver age, caregiver education, adversity score, children’s books at home). Model 3 is adjusted for trial factors from Model 1 and early-life factors (length for age Z-score (LAZ) at 18mo, birthweight, maternal baseline depression score (EPDS), household diet, maternal haemoglobin, socioeconomic status, facility birth, gender norms, and maternal years of schooling). Overall, 25 Plus-EF measurements were missing due to a programming error on encryption which led to some results being lost. For fine motor assessments, 6 children did not perform the task: 1 child’s caregiver refused, and 5 children were unable to fully understand or concentrate for the finger tapping task. Two caregivers did not answer all SDQ questions. Overall, 25 children refused to answer all questions on the child socioemotional scale, hence were unable to provide a full score.

| Outcome | CBHF |  | CHU |  | GEE Mean difference (95% CI) |  |  |  |
| --- | --- | --- | --- | --- | --- | --- | --- | --- |
|  | N | Mean (SD) | N | Mean (SD) | Unadjusted difference | Adjusted difference Model 1 (Trial factors) | Adjusted difference Model 2 (Trial factors & contemporary covariates) | Adjusted difference Model 3 (Trial factors & baseline covariates) |
| Mental Processing Index | 264 | 45 (11) | 990 | 48 (11) | 3 (2, 4) | 3 (2, 4) | 3 (1, 4) | 3 (2, 4) |
| School Achievement Test | 264 | 39 (26) | 990 | 46 (28) | 7 (4, 11) | 7 (3, 11) | 6 (2, 10) | 6 (3, 10) |
| Plus EF executive function score | 251 | 109 (25) | 978 | 114 (24) | 5 (2, 8) | 5 (2, 8) | 5 (2, 8) | 5 (2, 9) |
| Fine motor speed, seconds | 262 | 25.0 (6.7) | 986 | 24.1 (6.6) | -1.0 (-1.8, -0.1) | -0.8 (-1.7, 0.1) | -0.8 (-1.7, 0.1) | -0.7 (-1.6, 0.2) |
| Strengths & Difficulties Questionnaire score | 263 | 9 (5) | 989 | 9 (5) | 0 (-1, 0) | -1 (-1, 0) | 0 (-1, 1) | 0 (-1, 0) |
| Child socioemotional score | 256 | 4 (1) | 973 | 4 (1) | 0 (0, 0) | 0 (0, 0) | 0 (0, 0) | 0 (0, 0) |

Physical function scores for both groups are shown in Table 3. Whilst CBHF generally had lower scores for all tests, the only test with strong evidence of difference between groups was the level achieved in the shuttle run test, which reflects VO_2_max as a measure of cardiovascular fitness. CBHF compared to CHU had a 0.7 ml kg^-1^ min^-1^ lower VO_2_max, representing 0.3 lower average level (or approximately 60 metres shorter distance) on the shuttle run test. Growth and body composition for both groups are shown in Table 4. CBHF generally had lower growth and body composition values than CHU, but there was only strong evidence of difference in head circumference, which was 0.3 cm lower in CBHF, across all models. Taken together, CBHF generally had lower physical fitness scores and reduced growth, with strong evidence of difference for cardiovascular fitness and head circumference.

**Table 3:** Physical function in CBHF and CHU. Model 1 is adjusted for trial factors (arm, study nurse, exact child age, calendar month recruited, temperature, sex). Model 2 is adjusted for trial factors from Model 1 and contemporary factors (socioeconomic status, caregiver depression score (EPDS), household food insecurity (HFIAS), household religion, caregiver social support, caregiver gender norms, caregiver age, caregiver education, adversity score, children’s books at home). Model 3 is adjusted for trial factors from Model 1 and early-life factors (length for age Z-score (LAZ) at 18mo, birthweight, maternal baseline depression score (EPDS), household diet, maternal haemoglobin, socioeconomic status, facility birth, gender norms, and maternal years of schooling). Two children (both CBHF) did not perform the grip strength test: 1 was not motivated and 1 had a caregiver who refused the measurements. Eight children did not perform the broad jump test for a variety of reasons: 1 caregiver refused, 1 visit was performed during heavy rains and the ground was too slippery, 2 children had painful legs, 2 children’s caregivers refused due to the child being known to have asthma, 1 child refused and 1 measurement was missing without a recorded reason. In addition, 5 children did not do the shuttle run test: 3 children refused, 1 was recorded as having asthma, and 1 measurement was missing without a recorded reason. Two children refused blood pressure measurements.

| Outcome | CBHF |  | CHU |  | GEE Mean difference (95% CI) |  |  |  |
| --- | --- | --- | --- | --- | --- | --- | --- | --- |
|  | N | Mean (SD) | N | Mean (SD) | Unadjusted difference | Adjusted difference Model 1 (Trial factors) | Adjusted difference Model 2 (Trial factors & contemporary covariates) | Adjusted difference Model 3 (Trial factors & baseline covariates) |
| Grip Strength, Kg | 262 | 10.5 (1.9) | 990 | 10.7 (1.9) | 0.2 (-0.1, 0.5) | 0.3 (0.0, 0.5) | 0.3 (0.0, 0.5) | 0.2 (-0.0, 0.5) |
| Broad jump, cm | 259 | 111.0 (17.3) | 987 | 112.8 (15.1) | 2.1 (-0.2, 4.3) | 2.5 (0.3, 4.6) | 2.1 (-0.2, 4.3) | 1.8 (-0.3, 4.0) |
| VO2max, ml kg<sup>-1</sup> min<sup>-1</sup> | 255 | 50.2 (2.7) | 986 | 50.9 (2.7) | 0.8 (0.4, 1.2) | 0.6 (0.2, 1.0) | 0.5 (0.1, 0.9) | 0.5 (0.1, 0.9) |
| Systolic BP, mm Hg | 264 | 96.7 (9.0) | 988 | 97.0 (9.3) | 0.1 (-1.1, 1.2) | 0.5 (-0.5, 1.6) | 0.6 (-0.4, 1.7) | 0.6 (-0.5, 1.6) |
| Diastolic BP, mm Hg | 264 | 62.8 (7.3) | 988 | 62.3 (7.5) | -0.5 (-1.5, 0.4) | 0.0 (-0.9, 1.0) | 0.0 (-0.9, 1.0) | 0.1 (-0.9, 1.1) |

**Table 4:** Growth and body composition in CBHF and CHU. Model 1 is adjusted for trial factors (arm, study nurse, exact child age, calendar month recruited, temperature, sex). Model 2 is adjusted for trial factors from Model 1 and contemporary factors (socioeconomic status, caregiver depression score (EPDS), household food insecurity (HFIAS), household religion, caregiver social support, caregiver gender norms, caregiver age, caregiver education, adversity score, children’s books at home). Model 3 is adjusted for trial factors from Model 1 and early-life factors (length for age Z-score (LAZ) at 18mo, birthweight, maternal baseline depression score (EPDS), household diet, maternal haemoglobin, socioeconomic status, facility birth, gender norms, and maternal years of schooling). One caregiver refused all anthropometry measurements. Five children refused skinfold measurements, three children had a missing weight due to faulty weighing scales, nine children had missing bioimpedance measurements because of faulty machines or measurements that were excluded for inconsistency, two children refused haemoglobin measurements, and one child had missing knee-heel length, MUAC, waist and calf circumference measurements.

| Outcome | CBHF |  | CHU |  | GEE Mean difference (95% CI) |  |  |  |
| --- | --- | --- | --- | --- | --- | --- | --- | --- |
|  | N | Mean (SD) | N | Mean (SD) | Unadjusted difference | Adjusted difference Model 1 (Trial factors) | Adjusted difference Model 2 (Trial factors & contemporary covariates) | Adjusted difference Model 3 (Trial factors & baseline covariates) |
| Height-for-age Z score | 263 | -0.6 (0.9) | 990 | -0.5 (0.9) | 0.1 (0.0, 0.3) | 0.1 (0.0, 0.3) | 0.1 (0.0, 0.2) | 0.1 (0.0, 0.2) |
| Weight-for-age Z score | 262 | -0.7 (0.9) | 988 | -0.6 (0.9) | 0.1 (0.0, 0.2) | 0.1 (0.0, 0.2) | 0.1 (0.0, 0.2) | 0.1 (0.0, 0.2) |
| Body mass index, kg/m<sup>2</sup> | 262 | -0.5 (0.9) | 988 | -0.5 (0.8) | 0.0 (-0.2, 0.1) | 0.0 (-0.1, 0.1) | 0.0 (-0.2, 0.1) | 0.0 (-0.1, 0.1) |
| Knee-heel length, cm | 262 | 37.3 (2.0) | 989 | 37.4 (1.9) | 0.1 (-0.1, 0.4) | 0.2 (0.0, 0.5) | 0.2 (-0.1, 0.4) | 0.1 (-0.1, 0.4) |
| Head circumference, cm | 263 | 51.0 (1.5) | 990 | 51.3 (1.4) | 0.3 (0.1, 0.5) | 0.3 (0.1, 0.5) | 0.3 (0.1, 0.5) | 0.3 (0.1, 0.5) |
| Mid-upper arm circumference, cm | 262 | 17.0 (1.4) | 989 | 16.9 (1.3) | -0.1 (-0.3, 0.1) | 0.0 (-0.2, 0.2) | 0 (-0.2, 0.2) | 0.0 (-0.2, 0.1) |
| Waist circumference, cm | 263 | 54.1 (3.1) | 989 | 54.1 (3.1) | -0.1 (-0.6, 0.3) | 0.1 (-0.3, 0.5) | 0.1 (-0.3, 0.5) | 0.0 (-0.4, 0.4) |
| Hip circumference, cm | 263 | 61.0 (4.1) | 990 | 60.9 (3.9) | -0.1 (-0.7, 0.5) | 0.1 (-0.5, 0.7) | 0.1 (-0.5, 0.7) | 0.0 (-0.6, 0.6) |
| Calf circumference, cm | 263 | 23.3 (1.6) | 989 | 23.4 (1.7) | 0.1 (-0.1, 0.3) | 0.2 (0.0, 0.4) | 0.2 (0, 0.4) | 0.2 (-0.1, 0.4) |
| Lean mass index, Ohms<sup>-1</sup> | 262 | 12.1 (1.3) | 982 | 12.1 (1.3) | 0.0 (-0.2, 0.2) | 0.0 (-0.1, 0.2) | 0.0 (-0.2, 0.2) | 0.0 (-0.1, 0.2) |
| Impedance Index, m<sup>2</sup>/Ohms<sup>-1</sup> | 262 | 1.7 (0.3) | 982 | 1.8 (0.3) | 0.0 (0.0, 0.1) | 0.0 (0.0, 0.1) | 0.0 (0.0, 0.1) | 0.0 (0.0, 0.1) |
| Phase angle, degrees | 261 | 5.1 (0.5) | 982 | 4.9 (0.6) | -0.1 (-0.2, 0.0) | -0.1 (-0.1, 0.0) | -0.1 (-0.2, 0) | -0.1 (-0.1, 0.0) |
| Total skinfold thicknesses, mm | 261 | 26.4 (6.0) | 987 | 27.1 (6.2) | 0.6 (-0.3, 1.5) | 0.7 (-0.2, 1.6) | 0.7 (-0.2, 1.6) | 0.6 (-0.2, 1.5) |
| Peripheral skinfold thickness, mm | 261 | 15.7 (3.6) | 988 | 16.2 (3.7) | 0.5 (0.0, 1.0) | 0.5 (0.0, 1.0) | 0.5 (0.0, 1.0) | 0.5 (0.0, 1.0) |
| Central skinfold thickness, mm | 263 | 10.9 (3.1) | 989 | 10.9 (3.1) | 0.1 (-0.4, 0.5) | 0.1 (-0.3, 0.6) | 0.1 (-0.4, 0.7) | 0.1 (-0.4, 0.6) |
| Haemoglobin, g dl<sup>-1</sup> | 261 | 12.6 (1.3) | 990 | 12.7 (1.2) | 0.0 (-0.1, 0.2) | 0.1 (-0.1, 0.2) | 0.1 (-0.1, 0.3) | 0.1 (-0.1, 0.2) |

### Subgroup analysis

Among cognitive outcomes, there was evidence of an interaction between child sex and the Strengths and Difficulties questionnaire: among boys, CHU scored better than CBHF (1 mark, 95% CI 0, 2) but among girls, there was no evidence of difference in SDQ score between CBHF and CHU (Supplementary Table S3). Among physical function outcomes, there was evidence of an interaction between child sex and VO_2_max, whereby CHU boys had better cardiovascular fitness than CBHF boys (1.2, 95% CI 0.6, 1.8), while there was no evidence of difference between groups among girls. For growth outcomes, there was evidence of an interaction between sex and calf circumference such that CHU girls had weak evidence for a slightly larger calf circumference (0.3 cm, 95% CI 0.0, 0.6) but there was no evidence of a difference for boys. There were no significant interactions with child sex for other growth or body composition outcomes.

## Discussion

The population of children born HIV-free continues to expand due to improved coverage of PMTCT interventions among women living with HIV. It is now apparent that, despite ART during pregnancy, CBHF in sub-Saharan Africa have poorer early-life growth[7] and neurodevelopment[39]; however, few studies have evaluated child outcomes beyond 2 years of age and it remains unclear if disparities resolve, persist, or even widen over time. In this study from rural Zimbabwe, we show ongoing disparities in cognitive and physical function among CBHF at 7 years of age. Potential biological and environmental explanations for these disparities in CBHF are discussed below. The gap in neurodevelopment is substantial and interestingly is greater than the disparity in neurodevelopmental scores at age 2 years in the same cohort[20]. We also found that CBHF have reduced cardiovascular fitness, which may have consequences for adult health and function. Collectively, these data highlight the ongoing modest reductions in cognitive and physical function that CBHF experience, many years after exposure to HIV *in utero*, which may have a substantial long-term impact on human capital in areas of high HIV prevalence. There is an evident need to understand the underlying drivers of these differences, so that appropriate interventions can be deployed to improve long-term outcomes in this growing population of children.

The reduction in performance in CBHF was consistent across multiple domains of cognitive function including cognitive processing, academic and executive function. These tools had been specifically adapted[40] or developed for children at this age and in this setting[21]. CBHF compared to CHU had a 0.3 standard deviation reduction in the Mental Processing Index, representing the overall cognitive processing score, together with lower school achievement and executive function scores. This consistency across a range of cognitive domains has been previously shown in early life[8]. In a meta-analysis of 11 studies (6 outside the USA) CBHF showed reduced neurodevelopment compared to CHU at young ages, although sample sizes were small[39]. More recent studies have shown increased risk of language delay in CBHF[41, 42], but most studies are from children aged below 2 years. Here, we show that effects persist across cognitive domains by 7 years of age. Exploratory *post hoc* analysis showed reductions occurred across all subtests of these assessments, with the exception of the Flanker test in the executive function test battery (Supplementary Table S4). CBHF also had evidence of smaller head circumference, which has previously been noted in early life [43]. Reduced postnatal head circumference at age 2 years has been strongly associated with poor neurodevelopmental outcomes[44]. Evidence of structural differences among CBHF is emerging, with a reduction in grey matter volume apparent as early as 3 weeks of age using MRI[45], or diffuse tensor imaging combined with neuropsychological testing[46]. Head circumference is predominantly a marker of early-life growth, particularly in the first 2 years[47], and despite catch-up in other anthropometric measures over time, the signal of reduced head growth persists at school-age, consistent with the accompanying reductions in neurodevelopment.

It is likely that both universal and HIV-specific risk factors contribute to cognitive disparities[9]. The psychosocial and socioeconomic environment was poorer in CBHF, including worse food security, higher adversity scores, caregivers with fewer years of schooling, and higher caregiver depression scores, which may all affect the way caregivers provide nurturing care and how children learn. However, adjusted models including either contemporary or baseline psychosocial variables did not eliminate cognitive differences between groups, although the disparity was attenuated. Antenatal HIV, ART exposure, co-infections, and greater inflammation may drive biological differences in growth and function which are still evident at 7 years[16]. A combined intervention approach addressing both universal and HIV-specific pathways is likely needed to reduce the cognitive gap between groups, delivered throughout early life and up to school-age.

Cardiovascular fitness was also reduced in CBHF as shown by a reduction in the level obtained in the shuttle run, reflecting a lower VO_2_max than CHU. There could be several mechanisms for this. Emerging data suggest effects of HIV exposure on cardiac structure and function in high-income CBHF cohorts[48, 49]. Fetal exposure to ART may impair myocardial growth[49]. Zidovudine has been shown to alter fetal cardiac remodelling and to cause mild antenatal dysfunction[50]. It may also be due to HIV itself: In a large study of 400 American CBHF, greater cardiac inflammation and left ventricular stress were found in children living with HIV than those perinatally exposed to HIV[51]. A separate case-control study showed a 0.5 standard deviation reduction in left ventricular mass index in 30 school-age American CBHF compared to CHU[52]. A subgroup analysis in the current study showed that the reduction in cardiovascular fitness was only seen in boys. However, cardiac remodelling has been noted to be more pronounced in girls in the USA[48] so this sex difference may be due instead to differences in lung function between groups. CBHF have increased mortality from respiratory infections[9] which may also cause long-term morbidity: CBHF in South Africa showed negative associations between lung function and delays in starting ART, low maternal CD4 count and high viral loads[53]. Further detailed studies of cardiac and lung physiology are needed in cohorts of CBHF to better understand the reasons for reduced cardiovascular fitness in mid-childhood. The weak evidence for reduced calf circumference in CBHF girls did not associate with any evidence for reductions in strength or cardiovascular fitness for CBHF girls.

This study has several strengths, particularly the use of an extensively piloted toolbox[21] with suitably adapted assessments[23], which provided simultaneous phenotyping of growth, physical and cognitive function. The cohort has well-characterised longitudinal HIV exposure status, and includes a rich dataset including baseline and contemporary maternal, socioeconomic and nurturing factors for use as covariates. This substudy is likely to be representative of the broader SHINE cohort since there were no significant differences between those included and excluded in the follow-up study. There are also several study limitations, particularly possible survivor bias due to higher mortality in CBHF[7]. There may also be selection bias due to the high number of relocations since the end of the trial, which may be related to household socioeconomic status. Although we collected data during pregnancy on HIV treatment, it was not available for all women, and this substudy was not powered to evaluate the impact of specific antiretroviral regimens on long-term child health outcomes. Finally, we measured several outcomes and did not adjust for multiple comparisons since this was an exploratory analysis with many correlated outcomes; however, the consistent findings across cognitive tests provides reassurance that these results are unlikely simply due to inflated type 1 error.

In conclusion, this study of CBHF and CHU in rural Zimbabwe represents one of the few birth cohorts in sub-Saharan Africa to be followed to school-age to ascertain longer-term outcomes. We show ongoing vulnerabilities among CBHF in multiple domains of cognitive function and cardiovascular fitness, which could affect health and human capital across the life-course. The pressing goal now is to understand the relative contributions of biological and psychosocial factors to these long-term disparities, to inform future interventions, which could potentially include nurturing care and educational provision for children, combined with improved food security and psychosocial support to caregivers[16]. Given the expanding global population of CBHF there is a need for further long-term follow-up studies to understand how we can ensure that all children survive and thrive across the life-course.

## Data Availability

Data will be freely available as individual participant data on ClinEpiDB with an accompanying data dictionary at http://ClinEpiDB.org from 2025. Researchers must agree to the policies and comply with the mechanism of ClinepiDB to access data housed on this platform. Prior to that time, data are available upon reasonable request from the Zvitambo Institute for Maternal and Child Health Research, by contacting Dr Robert Ntozini

https://osf.io/8e2zh

## Supporting information: Growth, physical and cognitive function in children who are born HIV-free: school-age follow-up of a cluster-randomized trial in rural Zimbabwe

### Further description of scales used in baseline questionnaire

The socioeconomic status was recorded using a locally validated wealth index based on the household’s type of buildings and ownership of assets such as a wheelbarrow or chickens[1], with a higher score indicating greater household wealth. The coping strategies index is a validated measure of food insecurity, with a higher score showing greater food insecurity[2]. Caregiver depression was measured using the Edinburgh Postnatal Depression Scale, a 10-item self-report questionnaire which has previously been validated as a screening tool for depression in Zimbabwe[3], where a higher score represents more severe depressive symptoms. A higher score in gender norm attitudes reflects more equitable gender norms in the caregiver[4]. Similarly, an increase in the social support score reflects the caregiver’s perception of improved support from neighbours and family, particularly in sharing problems[4]. Both gender norms and social support scores have previously been associated with improved child growth at 18 months [5]. Maternal schooling exposure was measured by number of years of education.

### Further description of scales used in contemporary caregiver questionnaire

Child schooling exposure was measured in number of years of education combined with months of school attended in the current year. Adversity was measured using a range of questions exploring recent adversities in the child, caregiver or household, based on previous work in India[6] and used in a pilot study[7], with a higher score reflecting greater adversity. Nurturing was measured using the Child-Parent Relationship scale, where a higher score represents a more positive relationship of the child with the caregiver[8]. The MICS Child Discipline questionnaire asks a series of questions on methods of discipline used for the child, with a higher score reflecting more harsh methods of discipline[9, 10]. The Household Food Insecurity Assessment Scale (HFIAS) reflects food coping behaviours in the previous 28 days, with a higher score indicating greater food insecurity[11]. The Household Dietary Diversity Scale (HDDS) represents the range of foods eaten in the past week, with a higher score representing greater diversity[12]. Both HFIAS and HDDS have been previously used in Zimbabwe together to describe food security[13]. The Household Water Insecurity Experiences Scale (HWISE) also reflects water scarcity coping behaviours within the past 28 days, with a higher score representing more water insecurity[14].

**Table S1a.** Baseline characteristics of HIV-positive women and their infants enrolled or not enrolled in the follow-up cohort.

|  | Included in follow-up | Not included in follow-up | p-value |
| --- | --- | --- | --- |
| Caregiver assessed, N* | 267 | 459 |  |
| Children assessed, N | 273 | 465 |  |
| **Household characteristics** |  |  |  |
| Median number of occupants [IQR] | 4.0 (3.0 ; 6.0) | 4.0 (3.0 ; 6.0) | 0.03 |
| Wealth quintile, n (%) |  |  |  |
| Lowest | 64/258 (24.8%) | 129/453 (28.5%) | 0.49 |
| Second | 55/258 (21.3%) | 108/453 (23.8%) |  |
| Middle | 57/258 (22.1%) | 83/453 (18.3%) |  |
| Fourth | 41/258 (15.9%) | 63/453 (13.9%) |  |
| Highest | 41/258 (15.9%) | 70/453 (15.5%) |  |
| **Electricity** |  |  |  |
| Electricity in home, n (%) | 6/260 (2.3%) | 13/452 (2.9%) | 0.65 |
| Other electric power, n (%) |  |  |  |
| Generator | 9/260 (3.5%) | 9/453 (2.0%) | 0.10 |
| Solar panel | 171/260 (65.8%) | 269/453 (59.4%) |  |
| Inverter | 5/260 (1.9%) | 8/453 (1.8%) |  |
| No other type | 75/260 (28.9%) | 167/453 (36.9%) |  |
| **Sanitation** |  |  |  |
| Any latrine at household, n (%) | 92/255 (36.1%) | 142/447 (31.8%) | 0.28 |
| Improved latrine at household, n (%) | 79/255 (31.0%) | 124/446 (27.8%) | 0.42 |
| **Water** |  |  |  |
| Main source of household drinking water improved, n (%) | 159/255 (62.4%) | 263/446 (59.0%) | 0.46 |
| Treat drinking water to make it safer, n (%) | 28/255 (11.0%) | 56/438 (12.8%) | 0.50 |
| One-way walk time to fetch drinking water (min) , median (IQR) | 10.0 (5.0 ; 25.0) | 10.0 (5.0 ; 20.0) | 0.01 |
| Per capita water volume collected past 24 hr, median (IQR) | 6.7 (4.0 ; 10.0) | 8.6 (5.0 ; 13.3) | <0.001 |
| **Hygiene** |  |  |  |
| Handwashing station at household, n (%) | 35/246 (14.2%) | 33/412 (8.0%) | 0.02 |
| Improved floor, n (%) | 115/258 (44.6%) | 222/444 (50.0%) | 0.20 |
| Number of chickens, median (IQR) | 5.0 (2.0 ; 10.0) | 4.0 (0.0 ; 8.0) | 0.01 |
| Livestock observed inside the house, n (%) | 103/258 (39.9%) | 140/453 (30.0%) | 0.02 |
| Faeces observed in the yard, n (%) | 86/256 (33.6%) | 123/450 (27.3%) | 0.09 |
| **Diet quality and food security** |  |  |  |
| Household meets minimum dietary diversity score, n (%) | 85/243 (35.0%) | 151/369 (40.9%) | 0.15 |
| Coping strategies index, median (IQR) | 3.0 (0.0 ; 9.0) | 2.0 (0.0 ; 12.0) | 0.12 |
| **Maternal characteristics** |  |  |  |
| Mean age (SD), years | 30.2 (6.1) | 28.6 (6.3) | <0.001 |
| Mean height (SD), cm | 160.2 (6.2) | 160.2 (6.2) | 0.88 |
| Mean mid-upper-arm circumference (SD), cm | 26.5 (2.8) | 26.1 (3.00) | 0.01 |
| Mean maternal Hb (SD), g/dL | 11.5 (1.4) | 11.1 (1.8) | 0.01 |
| Mother meets minimum dietary diversity score, n (%) | 90/250 (36.0%) | 182/439 (41.5%) | 0.16 |
| Mean years of schooling completed (SD) | 9.2 (1.9) | 9.1 (2.2) | 0.54 |
| Median parity (IQR) | 2.0 (1.0 ; 3.0) | 2.0 (1.0 ; 3.0) | 0.01 |
| Married, n (%) | 231/248 (93.2%) | 412/434 (94.9%) | 0.71 |
| Employed, n (%) | 17/260 (6.5%) | 50/450 (11.1%) | 0.04 |
| Religion, n (%) |  |  |  |
| Apostolic | 118/252 (46.8%) | 212/437 (48.5%) | 0.85 |
| Other Christian | 106/252 (42.1%) | 182/437 (41.7%) |  |
| Other religion | 28/252 (11.1%) | 43/437 (9.8%) |  |
| Maternal capabilities |  |  |  |
| Median Gender norms attitudes (IQR) | 1.8 (1.5 ; 3.0) | 1.7 (1.5 ; 3.0) | 0.57 |
| Median Perceived social support (IQR) | 3.5 (3.0 ; 3.9) | 3.5 (3.1 ; 4.0) | 0.04 |
| Median EPDS depression scale (IQR) | 2.0 (0.0 ; 8.0) | 2.0 (0.0 ; 6.0) | 0.01 |
| **Infant characteristics** |  |  |  |
| Female, n (%) | 137/273 (50.2%) | 230/460 (50.0%) | 0.96 |
| Mean birth weight (SD), kg | 3.0 (0.5) | 3.0 (0.5) | 0.01 |
| Low birthweight, n (%) | 26/254 (10.2%) | 58/397 (14.6%) | 0.10 |
| Institutional delivery, n (%) | 216/247 (87.5%) | 328/402 (81.6%) | 0.06 |
| Vaginal delivery, n (%) | 240/258 (93.0%) | 369/401 (92.0%) | 0.73 |
| **18 month endpoint characteristics** |  |  |  |
| Mean LAZ at 18 months, (SD) | -1.9 (1.1) | -1.9 (1.2) | 0.92 |
| Mean WAZ at 18 months, (SD) | -1.0 (1.1) | -0.9 (1.1) | 0.51 |
| Mean WHZ at 18 months, (SD) | -0.1 (1.1) | -0.02 (1.2) | 0.31 |
| Mean Head circumference Z score at 18 months, (SD) | -0.5 (1.1) | -0.5 (1.2) | 0.90 |
| Mean MUAC Z score at 18 months, cm (SD) | -0.1 (0.9) | -0.2 (0.9) | 0.01 |
| Mean Hb at 18 months, g/dL (SD) | 11.8 (1.2) | 11.8 (1.2) | 0.79 |

**Table S1b.** Baseline characteristics of HIV-negative women and their infants enrolled or not enrolled in the follow-up cohort.

| **Supplementary table 1** | **HUU included in SFU (1002)** | **HUU Not included in SFU** | **p-value** |
| --- | --- | --- | --- |
| Caregiver assessed, N* | 988 | 2949 |  |
| Children assessed, N | 1002** | 2987 |  |
| **Household characteristics** |  |  |  |
| Median number of occupants [IQR] | 5.0 (4.0; 6.0) | 5.5 (3.0 ; 6.0) | 0.09 |
| Wealth quintile, n (%) |  |  |  |
| Lowest | 164/914 (17.9%) | 511/2744 (18.6%) | 0.84 |
| Second | 168/914 (18.4%) | 544/2744 (19.8%) |  |
| Middle | 186/914 (20.4%) | 553/2744 (20.2%) |  |
| Fourth | 200/914 (21.9%) | 576/2744 (21.0%) |  |
| Highest | 196/914 (21.4%) | 560/2744 (20.4%) |  |
| **Electricity** |  |  |  |
| Electricity in home, n (%) | 31/914 (3.4%) | 69/2738 (2.5%) | 0.18 |
| Other electric power, n (%) |  |  |  |
| Generator | 32/914 (3.5%) | 86/2743 (3.1%) | 0.21 |
| Solar panel | 631/914 (69.0%) | 1795/2743 (65.4%) |  |
| Inverter | 13/914 (1.4%) | 44/ 2743 (1.6%) |  |
| No other type | 238/914 (26.0%) | 818/2743 (29.8%) |  |
| **Sanitation** |  |  |  |
| Any latrine at household, n (%) | 343/899 (38.2%) | 981/2707 (36.2%) | 0.42 |
| Improved latrine at household, n (%) | 290/898 (32.3%) | 867/2703 (32.1%) | 0.92 |
| **Water** |  |  |  |
| Main source of household drinking water improved, n (%) | 618/900 (68.7%) | 1675/2725 (61.5%) | 0.01 |
| Treat drinking water to make it safer, n (%) | 127/895 (14.2%) | 322/2673 (12.1%) | 0.12 |
| One-way walk time to fetch drinking water (min) , median (IQR) | 10.0 (5.0 ; 20.0) | 10.0 (5.0 ; 20.0) | 0.30 |
| Per capita water volume collected past 24 hr, median (IQR) | 6.7 (4.4 ; 10.0) | 7.5 (5.0 ; 12.0) | <0.001 |
| **Hygiene** |  |  |  |
| Handwashing station at household, n (%) | 101/876 (11.5%) | 207/2557 (8.1%) | 0.01 |
| Improved floor, n (%) | 484/900 (53.8%) | 1512/2707 (55.9%) | 0.44 |
| Number of chickens, median (IQR) | 6.0 (2.0 ; 10.0) | 6.0 (2.0 ; 10.0) | 0.12 |
| Livestock observed inside the house, n (%) | 390/963 (40.5%) | 1039/2886 (36.0%) | 0.02 |
| Faeces observed in the yard, n (%) | 322/958 (33.6%) | 877/2879 (30.5%) | 0.15 |
| **Diet quality and food security** |  |  |  |
| Household meets minimum dietary diversity score, n (%) | 328/880 (37.3%) | 965/2344 (41.2%) | 0.10 |
| Coping strategies index, median (IQR) | 0.0 (0.0 ; 6.0) | 1.0 (0.0 ; 7.0) | 0.01 |
| **Maternal characteristics** |  |  |  |
| Mean age (SD), years | 25.7 (6.3) | 25.6 (6.7) | 0.15 |
| Mean height (SD), cm | 159.9 (6.0) | 160.2 (5.6) | 0.08 |
| Mean mid-upper-arm circumference (SD), cm | 26.6 (3.3) | 26.3 (3.0) | 0.05 |
| Maternal Hb, g/dL (SD) | 12.2 (1.4) | 12.1 (1.5) | 0.21 |
| Mother meets minimum dietary diversity score, n (%) | 330/901 (36.6%) | 1081/2675 (40.4%) | 0.10 |
| Mean years of schooling completed (SD) | 9.7 (1.7) | 9.6 (1.8) | 0.10 |
| Median parity (IQR) | 2.0 (1.0 ; 3.0) | 2.0 (1.0 ; 3.0) | 0.03 |
| Married, n (%) | 889/935 (95.1%) | 2657/2782 (95.5%) | 0.29 |
| Employed, n (%) | 69/916 (7.5%) | 242/2739 (8.8%) | 0.23 |
| Religion, n (%) |  |  |  |
| Apostolic | 455/941 (48.4%) | 1308/2804 (46.7%) | 0.097 |
| Other Christian | 395/941 (42.0%) | 1289/2804 (46.0%) |  |
| Other religion | 91/941 (9.7%) | 207/2804 (7.4%) |  |
| Maternal capabilities |  |  |  |
| Median Gender norms and attitudes (IQR) | 2.3 (1.5 ; 3.0) | 1.7 (1.5 ; 3.0) | <0.001 |
| Median Perceived social support (IQR) | 3.6 (3.2 ; 4.0) | 3.7 (3.3 ; 4.0) | 0.05 |
| Median EPDS depression scale (IQR) | 1.0 (0.0 ; 5.0) | 1.0 (0.0 ; 4.0) | 0.81 |
| **Infant characteristics** |  |  |  |
| Female, n (%) | 511/1002 (51.0%) | 1451/2972 (48.8%) | 0.28 |
| Mean birth weight (SD), kg | 3.1 (0.5) | 3.1 (0.5) | 0.43 |
| Low birthweight, n (%) | 90/957 (9.4%) | 235/2616 (9.0%) | 0.74 |
| Institutional delivery, n (%) | 855/935 (91.4%) | 2353/2669 (88.2%) | 0.01 |
| Vaginal delivery, n (%) | 909/965 (94.2%) | 2482/2699 (92.0%) | 0.10 |
| **18 month endpoint characteristics** |  |  |  |
| Mean LAZ at 18 months, (SD) | -1.5 (1.0) | -1.5 (1.1) | 0.18 |
| Mean WAZ at 18 months, (SD) | -0.7 (1.0) | -0.7 (1.0) | 0.06 |
| Mean WHZ at 18 months, (SD) | -0.04 (1.0) | 0.1 (1.1) | 0.001 |
| Mean Head circumference Z score at 18 months, (SD) | -0.2 (1.1) | -0.2 (1.5) | 0.48 |
| Mean MUAC Z score at 18 months (SD) | 0.1 (0.9) | 0.02 (0.9) | 0.03 |
| Mean Hb at 18 months, g/dL (SD) | 11.8 (1.1) | 11.7 (1.2) | 0.12 |

**Table S2:** Baseline characteristics of children born HIV-free (CBHF) and children HIV-unexposed (CHU)

| **Maternal characteristics in pregnancy** | **CBHF** | **CHU** | **p-value** |
| --- | --- | --- | --- |
| Mean age (SD), years | 30.3 (6.0) | 25.7 (6.3) | <0.001 |
| Mean height (SD), cm | 160.4 (6.1) | 160.0 (6.0) | 0.35 |
| Mean MUAC (SD), cm | 26.6 (2.8) | 26.6 (3.2) | 0.99 |
| Mean haemoglobin (SD), g/dl | 11.5 (1.4) | 12.2 (1.4) | <0.001 |
| Mean years of schooling (SD) | 9.1 (1.9) | 9.7 (1.7) | <0.001 |
| Median parity (IQR) | 2 (1, 3) | 2 (1, 3) | <0.001 |
| Married, % [N] | 93.5% [229/245] | 95.2% [893/938] | 0.45 |
| Employed, % [N] | 6.6% [17/256] | 7.5% [69/919] | 0.64 |
| Religion, % [N] |  |  |  |
| Apostolic, % [N] | 45.6% [113] | 48.1% [454] | 0.60 |
| Other Christian | 42.7% [106] | 42.0% [398] |  |
| Other non-Christian | 11.7% [29] | 9.8% [92] |  |
| Maternal capabilities |  |  |  |
| Mean Gender norms (SD) | 2.2 (0.8) | 2.3 (0.8) | 0.16 |
| Mean perceived social support (SD) | 3.4 (0.7) | 3.5 (0.6) | 0.03 |
| Mean Edinburgh Postnatal Depression Score (SD) | 4.6 (5.6) | 2.8 (4.0) | <0.001 |
| **Maternal HIV disease severity and treatment** |  |  |  |
| Mean CD4 count in pregnancy, cells/uL (SD) | 474.3 (SD 214.6) [n=210] | N/A |  |
| Documented antiretroviral therapy during pregnancy | 82.8% [226/273] | N/A |  |
| Tenofovir disoproxil fumarate-based ART regimen | 69.9% [158/226] | N/A |  |
| Zidovudine based ART regimen | 18.6% [42/226] | N/A |  |
| Other/unknown ART regimen | 11.5% [26/226] | N/A |  |
| Documented co-trimoxazole prophylaxis during pregnancy | 66.7% [182/273] | N/A |  |
| **Household characteristics in pregnancy** |  |  |  |
| Median household size (IQR) | 4 (3, 6) | 5 (4, 6) | 0.08 |
| Median Coping Strategies Index (IQR) | 3 (0, 9) | 0 (0, 9) | <0.001 |
| Baseline wealth quintile, % [N] |  |  |  |
| lowest | 25.2% [64 /254] | 17.7% [162 / 917] | 0.007 |
| second | 21.7% [55 / 254] | 18.4% [169 / 917] |  |
| middle | 21.7% [55 / 254] | 20.5% [188 / 917] |  |
| fourth | 15.0% [38 / 254] | 21.7% [199 / 917] |  |
| highest | 16.5% [42 / 254] | 21.7% [199 / 917] |  |
| Electricity in house, % [N] | 2.3% [6 / 256] | 3.4% [31 / 917] | 0.40 |
| Any latrine, % [N] | 36.3% [91 / 251] | 38.0% [343 / 902] | 0.61 |
| Improved latrine, % [N] | 30.7% [77 / 251] | 32.1% [289 / 901] | 0.67 |
| Improved water source, % [N] | 61.8% [155 / 251] | 68.3% [617 / 903] | 0.05 |
| Treat drinking water in any way, % [N] | 11.2% [28 / 251] | 14.5% [130 / 898] | 0.18 |
| HH meets min dietary diversity, % [N] | 36.4% [87 / 239] | 37.7% [333 / 884] | 0.72 |
| Women meet min dietary diversity score, % [N] | 37.4% [92 / 246] | 36.8% [333 / 904] | 0.87 |
| **Child Variables** | **CBHF** | **CHU** | **p-value** |
| Mean Birthweight, Kg (SD) | 3.03 (0.46) | 3.10 (0.47) | 0.06 |
| Proportion Low Birthweight, % [N] | 10.2% [25 / 246] | 9.2% s[87 / 945] | 0.65 |
| Proportion Institutional delivery, % [N] | 87.0% [207 / 238] | 91.4% [844 / 923] | 0.04 |
| Proportion vaginal delivery, % [N] | 93.2% [232 / 249] | 94.4% [900 / 953] | 0.45 |
| Mean breastfeeding duration, months (SD) | 17.7 (4.5) | 19.1 (3.7) | <0.001 |
| Mean 18-month LAZ (SD) | -1.83 (1.06) | -1.50 (1.02) | <0.001 |
| Stunted at 18 months, % [N] | 45.2% [118 / 261] | 29.6% [290 / 981] | <0.001 |
| Mean 18-month WAZ (SD) | -0.96 (1.05) | -0.74 (0.98) | 0.003 |
| Underweight at 18 months, % [N] | 16.9% [44 / 261] | 8.8% [86 / 981] | <0.001 |
| Mean 18-month WHZ (SD) | -0.11 (1.06) | -0.03 (1.00) | 0.28 |
| Mean 18-month HCZ (SD) | -0.46 (1.07) | -0.21 (0.98) | 0.001 |
| Mean 18-month MUACZ (SD) | -0.06 (0.87) | 0.10 (0.87) | 0.01 |
| Mean 18-month Hb, g/dL (SD) | 11.8 (1.1) | 11.8 (1.1) | 0.93 |

### Directed Acyclic Graph for Contemporary Covariates

**Figure 0-1:**
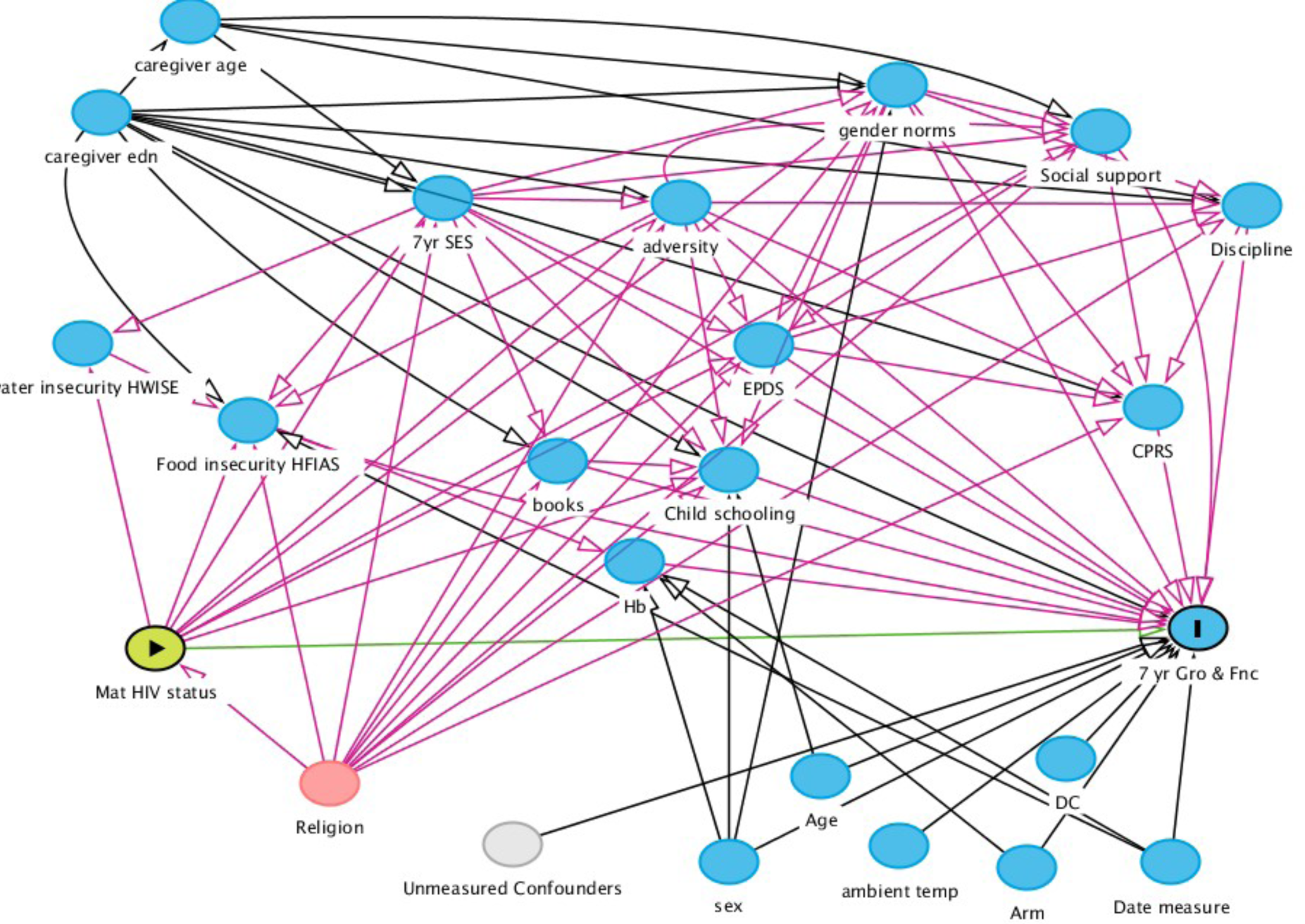
Directed Acyclic Graph (DAG) used to determine covariates for Model 2: adjustment by contemporary covariates asked in the contemporary questionnaire. DC: Data collector, sex: Child sex, Arm: SHINE trial intervention arm, Date measure: calendar quarter when measurement performed, Exact age: exact age of child, ambient temperature: average temperature during SAHARAN toolbox measurements, Confounders: unmeasured confounders, Caregiver edn: Caregiver schooling in number of years, Caregiver age: age of primary caregiver at 7 year visit, 7yr SES: contemporary socioeconomic status (wealth index), adversity: contemporary adversity score,, EPDS: contemporary caregiver Edinburgh Postnatal Depression Score, Gender norms: contemporary caregiver gender norm scale, Social support: contemporary caregiver social support scale, Water insecurity (HWISE): Household water insecurity experiences scale, Food insecurity (HFIAS): Household food insecurity experiences scale, Books: number of children’s books at home, Child schooling: Total child schooling in years and months, Hb: child contemporary haemoglobin measured during visit, CPRS: Child parent relationship scale (measure of nurturing), Discipline: child discipline scale, Religion: household religion, mat HIV: Maternal HIV status during pregnancy (the exposure), 7yr Gro & fnc: child growth, cognitive and physical function at 7 years (the outcome). Adjustment variables for model 2 were arm, DC, age of child, calendar age recruited, temperature, sex, Socioeconomic status, Caregiver depression measure (EPDS), Household food insecurity (HFIAS), Household religion, Caregiver social support, Caregiver gender norms, Caregiver age, Caregiver education, Adversity score, Children’s books at home.

### Directed Acyclic Graph for Baseline Factors

**Figure 0-2:**
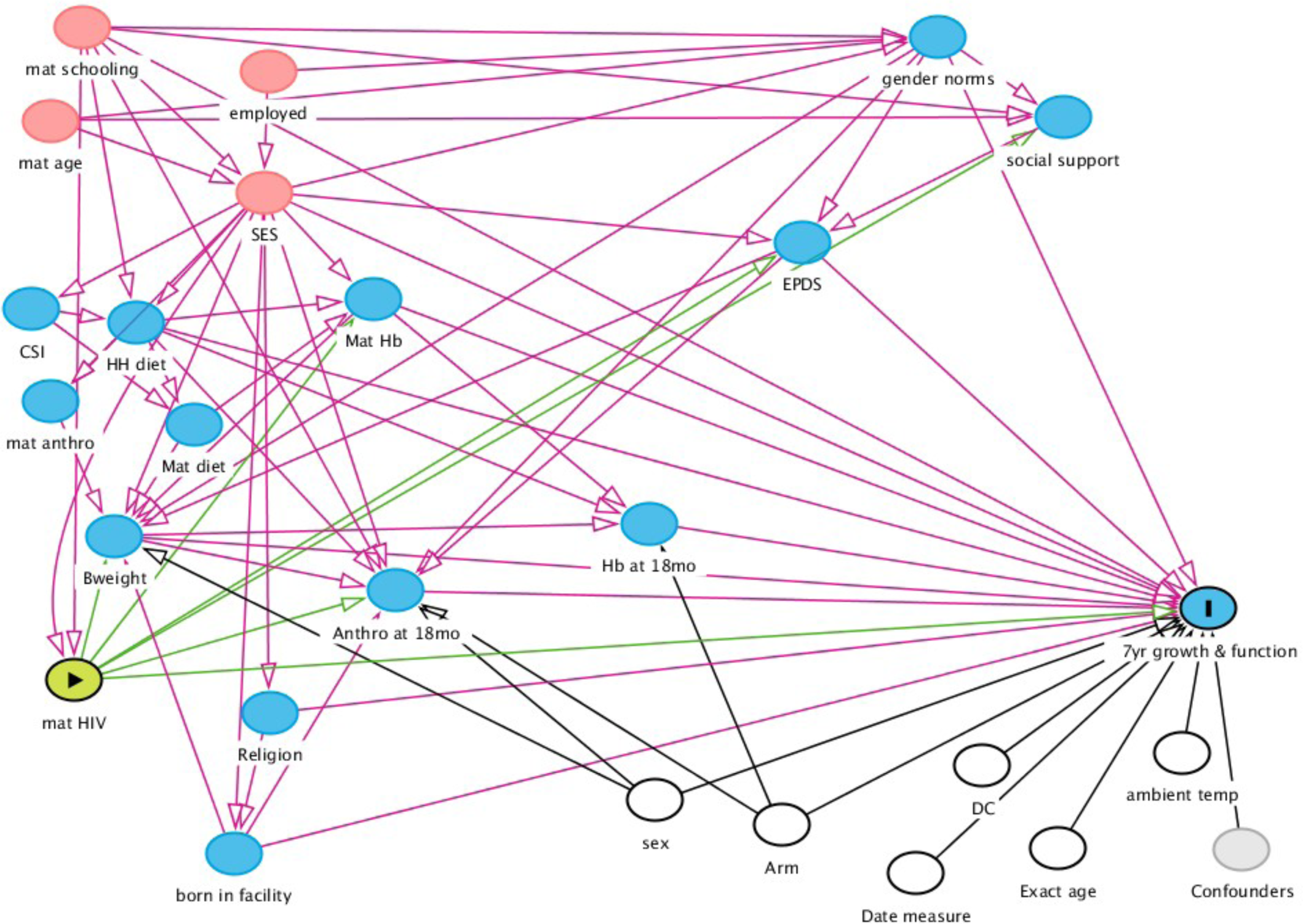
DAG used to determine covariates for Model 3: adjustment by baseline covariates. DC: Data collector, sex: Child sex, Arm: SHINE trial intervention arm, Date measure :calendar quarter when measurement performed, Exact age: exact age of child, ambient temperature: average temperature during SAHARAN toolbox measurements, Confounders: unmeasured confounders, Mat schooling: Maternal schooling in number of years, mat age: maternal age, SES: baseline socioeconomic status (wealth index), employed: whether mother was employed or not, EPDS: Baseline maternal Edinburgh postnatal depression score, Gender norms: baseline maternal gender norm scale, social support: baseline maternal social support scale, CSI: Coping strategies index (measure of food insecurity), HH diet: household dietary score, Mat HB: Maternal haemoglobin in pregnancy, Mat diet: maternal diet score, Mat anthro: Maternal anthropometry (note height was used in model), Bweight: child birthweight, mat HIV: Maternal HIV status during pregnancy (the exposure), anthro at 18 months: child anthropometry at 18 months (length-for-age-z-score used in model), religion: household religion, Hb at 18 months: child haemoglobin at 18 months of age. Adjustment variables were: Arm, cata collector, age of child, calendar age recruited, temperature, Anthropometry at 18mo, Birthweight, maternal depression score (EPDS), household dietary score, maternal haemoglobin in pregnancy, baseline socioeconomic scale, born in facility, gender norms, maternal years of schooling.

**S3.** Interaction analysis by child sex.

| Variable | P-value of interaction of HIV-exposure with child sex | GEE Coefficient for Girls (95% CI) | GEE coefficient for Boys (95% CI) |
| --- | --- | --- | --- |
| Mental Processing Index | 0.20 | N/A | N/A |
| School Achievement Test | 0.99 | N/A | N/A |
| Plus EF test score | 0.87 | N/A | N/A |
| Fine motor , sec | 0.58 | N/A | N/A |
| Strengths and Difficulties Questionnaire | 0.04 | 0.3 (-0.6, 1.2) | 1.2 (0.2, 2.3) |
| Child socioemotional score | 0.05 | 0.1 (-0.0, 0.2) | -0.1 (-0.2, 0.0) |
| Mean Grip Strength, Kg | 0.47 | N/A | N/A |
| Mean Broad jump, m | 0.61 | N/A | N/A |
| VO2max, ml kg<sup>-1</sup> min<sup>-1</sup> | 0.07 | 0.4 (-0.0, 0.8) | 1.1 (0.5, 1.7) |
| Diastolic BP, mm Hg | 0.89 | N/A | N/A |
| Systolic BP, mm Hg | 0.90 | N/A | N/A |
| Height-for-age Z-score | 0.37 | N/A | N/A |
| Weight-for-age Z-score | 0.99 | N/A | N/A |
| BMI Z-score | 0.30 | N/A | N/A |
| Knee-heel length | 0.63 | N/A | N/A |
| Head circ, cm | 0.72 | N/A | N/A |
| MUAC, cm | 0.24 | N/A | N/A |
| Waist circ, cm | 0.97 | N/A | N/A |
| Hip circ, cm | 0.37 | N/A | N/A |
| Calf circ, m | 0.08 | 0.3 (-0.0, 0.6) | -0.2 (-0.4, 0.2) |
| Lean mass index | 0.41 | N/A | N/A |
| Impedance Index | 0.84 | N/A | N/A |
| Phase angle | 0.45 | N/A | N/A |
| Total skinfold thicknesses, mm | 0.19 | N/A | N/A |
| Peripheral skinfold thickness, mm | 0.21 | N/A | N/A |
| Central skinfold thickness, mm | 0.29 | N/A | N/A |
| Hb, g dl<sup>-1</sup> | 0.30 | N/A | N/A |

**S4.**
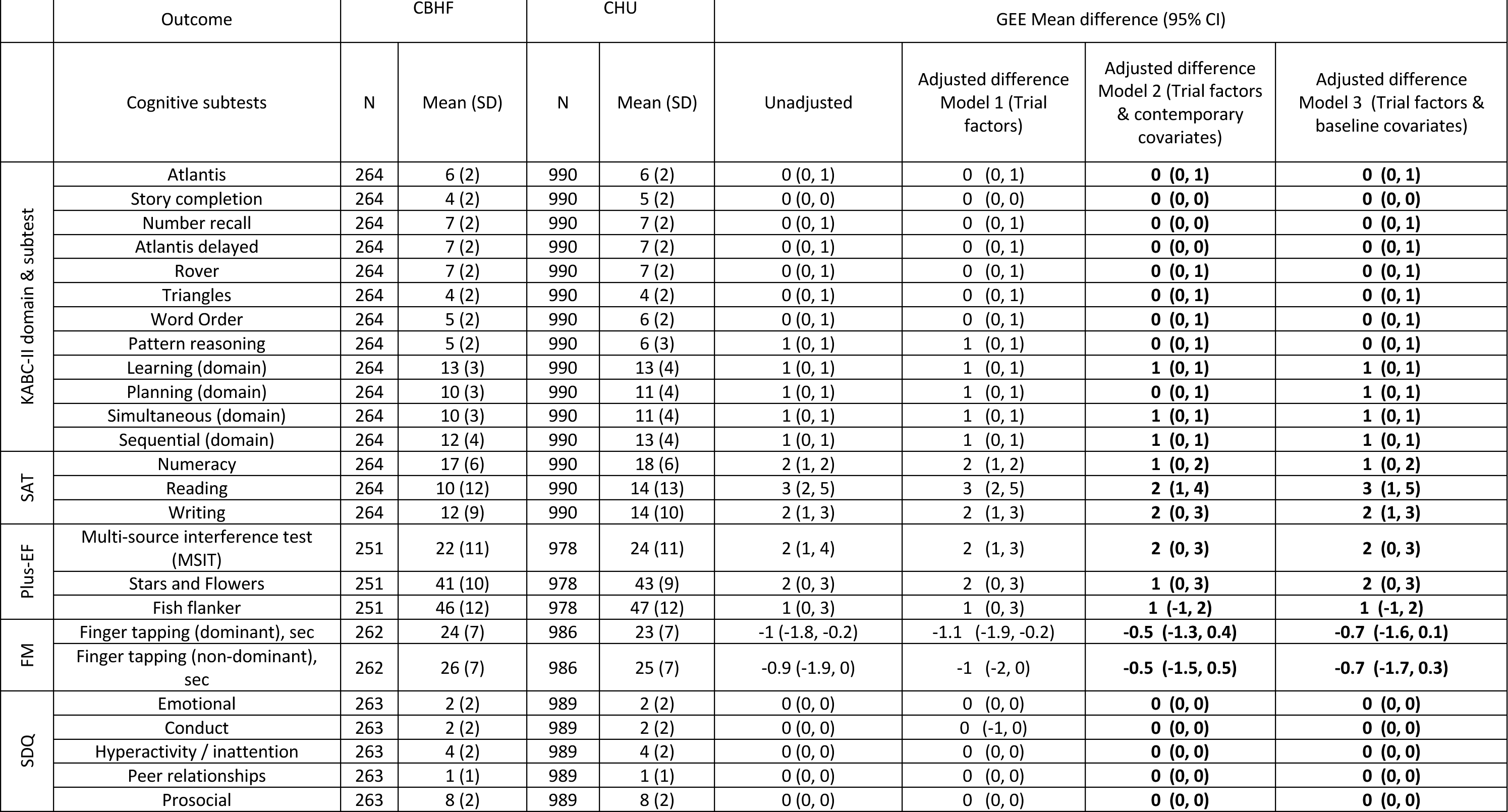
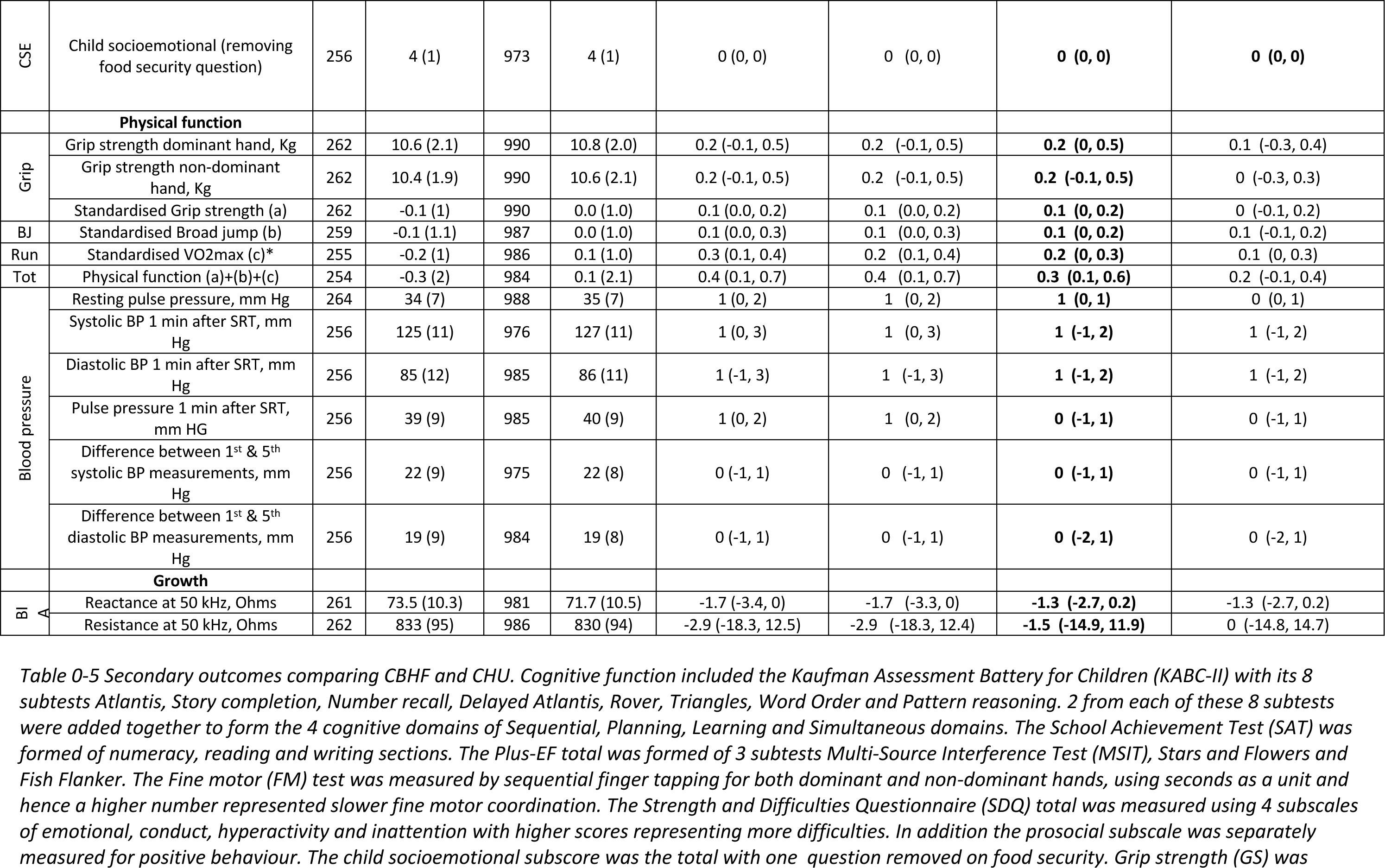
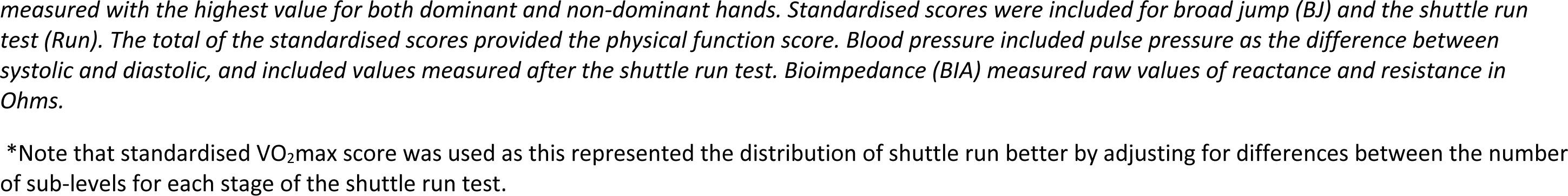
Cognition subtests & secondary physical function outcomes compared between CBHF and CHU.

### Description of secondary outcomes

The Kaufman Assessment Battery for Children 2^nd^ edition (KABC-II) assesses cognitive processing using Luria’s model by focusing on novel tests that have not been previously seen in schools. Hence the test results may be less dependent on overall schooling, which is suitable in circumstances where similar-aged children may have very different schooling exposure due to socioeconomic and environmental factors. The KABC-II can be condensed to eight subtests which are scaled based on the participant’s age, so that younger children have a higher scaled score for each test. The scaled results from two of the eight subtests are added together to create the four Luria domains of cognitive processing. The Sequential memory domain focuses on short-term sequential memory using the Number Recall and Word Order subtests. The Planning domain represents pattern recognition and problem-solving using the Story Completion and Pattern Reasoning subtests (this included the adaptations for Zimbabwe previously published[15]). Short- and long-term memory provide the Learning domain, measured using the Atlantis and Atlantis Delayed subtests. Finally, the Simultaneous domain measures logical and spatial problem solving using the Rover and Triangles subtests.

The school achievement test (SAT) was created previously from a pilot study cohort[7] and measured numeracy, reading and writing skills. Briefly, numeracy was the total from counting, visual and figure-based questions based on previous studies[16] [10]. Reading was the total from timed questions on reading letters, words and a local story, with participants given a choice of local languages (Shona, Ndebele or English) [16]. Writing was the total from the participant writing letters, words and their name in local languages[17].

The *PLUS-EF* is a tablet-based executive function tool, that has been used in school-aged children[18]. The score from multi-source interference (MSIT) was the total that reflected accuracy from the child picking the correct number from different sequences. Stars and Flowers and the Fish Flanker scores was the total in accuracy for the child picking the correct side for different shapes or arrows in different sequences.

Fine motor (FM) was the number of seconds to complete six repetitions of finger tapping in sequence from the thumb to the 5^th^ finger, divided into dominant and non-dominant hand, with a higher value indicating slower fine motor coordination.

The Strength and Difficulties questionnaire (SDQ) consists of 4 subscales with questions asking about emotional, behavioural, attention and relationship problems, with a higher score indicating more difficulties. These 4 subscales provided the SDQ total. A 5^th^ prosocial subscale asks about positive behaviours[19]. The child socioemotional (CSE) subscale removed the one question on food security, and remained with 5 questions on how the child felt they were supported at home.

For physical function, grip strength (GS) was subdivided into dominant and non-dominant hands, and a standardised score calculated. Broad jump (BJ) and shuttle run (Run) also had a standardised score calculated, so that a standardised physical function score was calculated. Blood pressure (BP) was measured at rest, where the pulse pressure was the difference between systolic and diastolic blood pressures. Blood pressure was also measured sequentially 5 times after the shuttle run test. All units were in mm Hg. Finally, raw units of reactance and resistance at 50 kHz were recorded for bioimpedance analysis (BIA).

